# Research report CERITER Clinical study - Stride One

**DOI:** 10.1101/2024.09.04.24312478

**Authors:** Sarah Meyer, Jan Limet, Piet Stevens, Marc Michielsen

## Abstract

**Objective:** This study investigates the short-term impact of the use of Ceriter Stride One on the quality of gait in patients rehabilitating after a cerebrovascular accident (CVA). Stride One is a smart insole that provides real-time audio feedback (cues) to the patient when performing a certain exercise. The goal of Stride One is to allow patients to practice walking more intensively and with higher quality during rehabilitation.

**Method:** Ten patients were offered therapy using Stride One for one week. The quality of the gait pattern was evaluated at the beginning and at the end of the week, with and without the use of Stride One. An additional evaluation was performed three to four days after the end of therapy without the use of Stride One. One patient was unable to complete the therapy due to a fall.

**Results:** Both therapist and patient reported an improvement in the quality of the foot roll-off pattern in 100% of cases. This improvement was also objectively determined in the data measured by Stride One, with an average of 8% improvement on Stride One general quality parameter, and an average of 25% improvement on Stride One specific quality parameter for the specific patient. Furthermore, 89% of patients indicate that using Stride One helps them to understand their physiotherapist better, and 67% of patients indicate that they can practice more and better with Stride One.

**Conclusion:** Practicing one week with Stride One generates a positive clinical impact on the gait pattern. A lasting improvement in the gait pattern without using Stride One, three to four days after the intervention, could not be demonstrated in this short time period. Further research is needed to evaluate the effect of Stride One after more long-term use.

Additional research questions are suggested.

## 1. Background

A cerebrovascular accident (CVA) is a multifactorial disorder that affects both cognitive and physical/motor parameters. One of the most important problems in rehabilitation after CVA (due to hemiplegia) is difficulty in walking, which leads to mobility problems and dependency of the patient on others. When walking, a patient with CVA shows a clear abnormality at the level of the foot, namely an abnormal roll-off pattern. This causes the patient to compensate in other parts of the body. However, if the roll-off pattern of the foot can be influenced or improved, the patient’s gait pattern will show an overall qualitative improvement. Ensuring a good foot roll-off is therefore a primary objective in gait rehabilitation after CVA.

Ceriter Stride One is a CE-certified medical device for qualitatively better and more independent walking. The device helps people with walking problems caused by neurological disorders such as Parkinson’s disease or CVA. Stride One detects a deviating gait pattern via a smart insole and generates individually tailored audio feedback, for example to prevent or correct a deviating roll-off pattern in the foot as a result of the CVA.

To date, no extensive clinical research has been conducted with Stride One in people with a CVA. In analogy with previously published scientific literature regarding the influence of audio feedback on the gait pattern in CVA and the intensity of exercise, it is expected that Stride One will ensure a better gait pattern in this patient population (see Bibliography 1-4). FRAME, the clinical test center of the Jessa Hospital (Belgium), was asked by CERITER to perform a Stride One clinical scientific assessment.

## 2. Objectives

The aim of this pilot study is to investigate whether patients with a CVA show a qualitatively better gait pattern after training with a Stride One insole, and whether patients can maintain this improvement in gait pattern without audio feedback at the end of the training.

Primary endpoints:

- Complete and correct foot roll-off (detected and interpreted in the accompanying software)
- Improved walking speed/greater number of steps per minute

Secondary endpoints:

- Improved functional tests: sit-to-stand, 3min walking test
- Subjective experiences: questionnaires

## 3. Method

### 3.1 Design

This pilot study consists of a case series (n=10) of patients with a CVA who received daily gait rehabilitation using Stride One for 1 week. As shown in the figure below, several measurements are performed before the start of the therapy (baseline measurement, T1), immediately after the end of the intervention period (post-intervention measurement, T2) and 2 days after the end of the therapy (follow-up measurement, T3).

**Figure 1.**
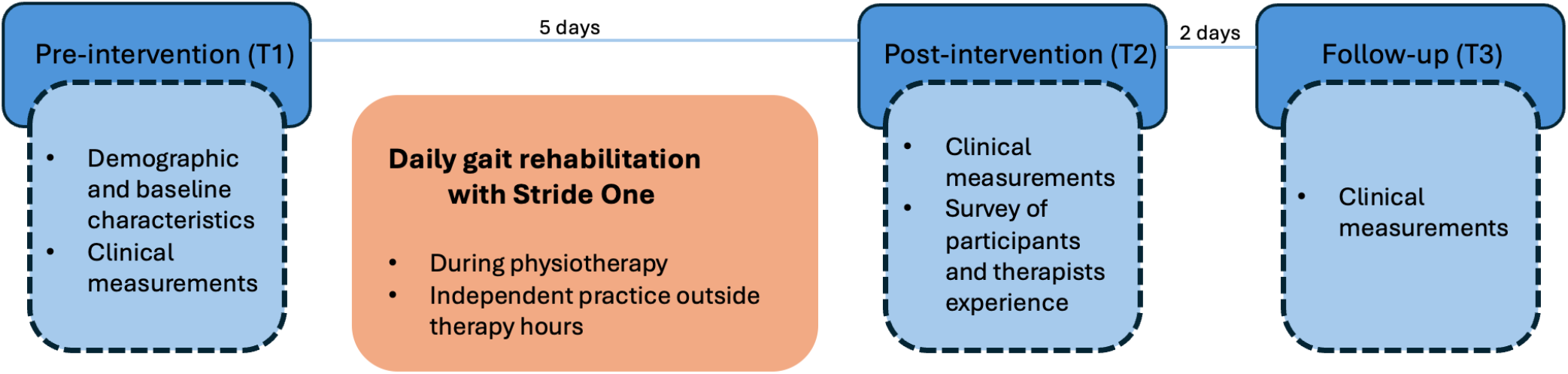
design.

### 3.2 Participants

In the context of this pilot study, 10 patients with a CVA were recruited between December 2023 and April 2024 from the rehabilitation center of the Jessa hospital (campus Sint-Ursula), using the following criteria:

Inclusion criteria:

– CVA, residentially admitted to the rehabilitation center in Herk-de-Stad, Belgium
– Older than 18 years of age
– Gait problems as a result of the CVA
– Able to walk safely and independently (possibly with a walking aid)
– Able to understand and sign an information and consent form

Exclusion criteria:

– Severe cognitive problems (attention) that make it difficult to understand instructions or follow feedback
– Hearing problem

The study was approved by the Jessa Hospital Medical Ethics Committee (2023/c/109).

### 3.3 Outcome measurements

The following demographic and baseline characteristics were collected at T1 (baseline):

- Year of birth
- Gender
- Date and affected side of CVA
- Date of independent walking
- Smoker
- Shoe size & possible use of orthopedic insoles
- Use of walking aid
- Use of own smartphone

The following clinical outcome measurements were performed at the 3 measurement moments (T1, T2 and T3):

- 5x sit-to-stand (seconds)
- 3-minute walking test without audio feedback (distance in meter + roll-off pattern via CERITER software)
- 3-minute walking test with audio feedback (distance in meter + roll-off pattern and recording of auditory feedback via CERITER software), only at T1 and T2

Immediately after the end of the intervention week (T2), the patient and the treating therapist were asked about their findings regarding the use of Stride One while walking.

After the study, a focus group was organized with all therapists involved to collect their insights, feedback and suggestions.

### 3.4 Intervention

During 1 week, daily therapy was offered to the participants, with Stride One. Stride One provides auditory feedback to the patient, stimulating the patient to achieve a good roll-off of the foot. During the support phase in the gait cycle, the roll-off pattern in the foot is crucial, and in a normal situation follows the steps below:

1. Heel-strike: heel first contacts the ground. This sends a signal to the central pattern generator (CPG) in the spinal cord to extend the hip and knee so that support can be achieved.
2. Mid-stance: the foot is rolled over the lateral side of the foot, towards the medial side and then to the big toe (c-shape)
3. Terminal stance: the foot is released from the ground via the big toe. Together with sufficient hip extension, this provides input to the CPG to flex the hip and knee towards the swing phase.

The aim of Stride One is to teach patients to place their feet correctly, in order to obtain a better roll-off, using positive audio feedback. During the first training, the feedback of the insole was personalized for each patient by the treating therapist. After the first training, the insole (with auditory feedback) was used daily in rehabilitation (physiotherapy), as well as by the patient themselves (outside therapy), and this for 5 days.

## 4. Results

### 4.1 Demographic data

The table below shows the demographic and baseline characteristics of the 10 participants in this pilot study. During the study, 1 drop-out (participant 6) was observed as a result of a fall during the gait rehabilitation training in the intervention week, resulting in a hip fracture. The study was stopped for this participant, so that no post-intervention (T2) and follow-up data (T3) are available.

**Table 1.**
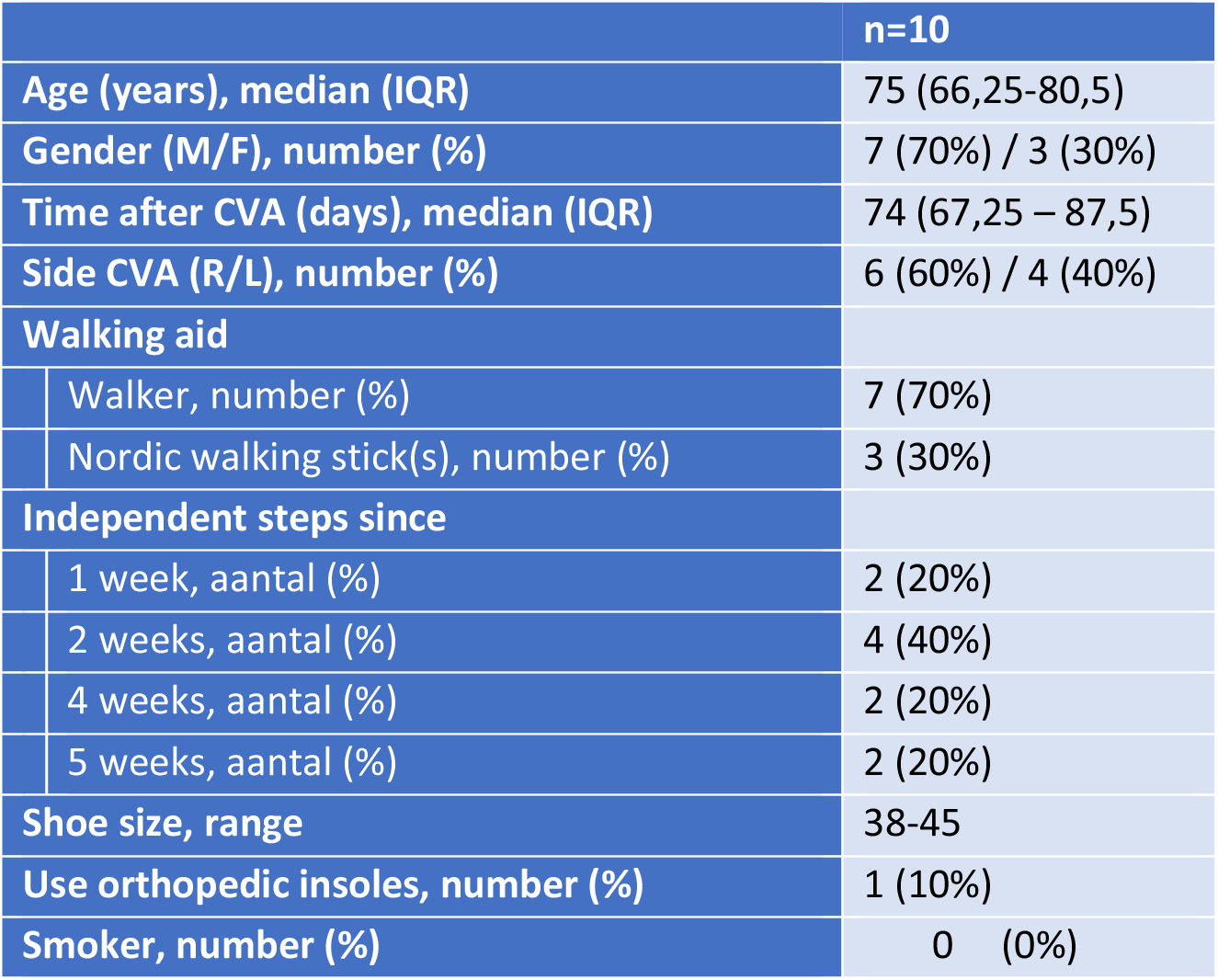
Demographic and baseline characteristics.

### 4.2 Clinical data

The following outcome measurements were performed at the 3 measurement moments (T1, T2 and T3):

- 5x sit-to-stand (seconds)
- 3-minute walk test without audio feedback (distance in meters, gait quality indicators were collected in Stride One)

The results of the manual measurements are shown in Figure 2 and Figure 3. Stride One measurements are covered in part 5 of the report.

**Figure 2.**
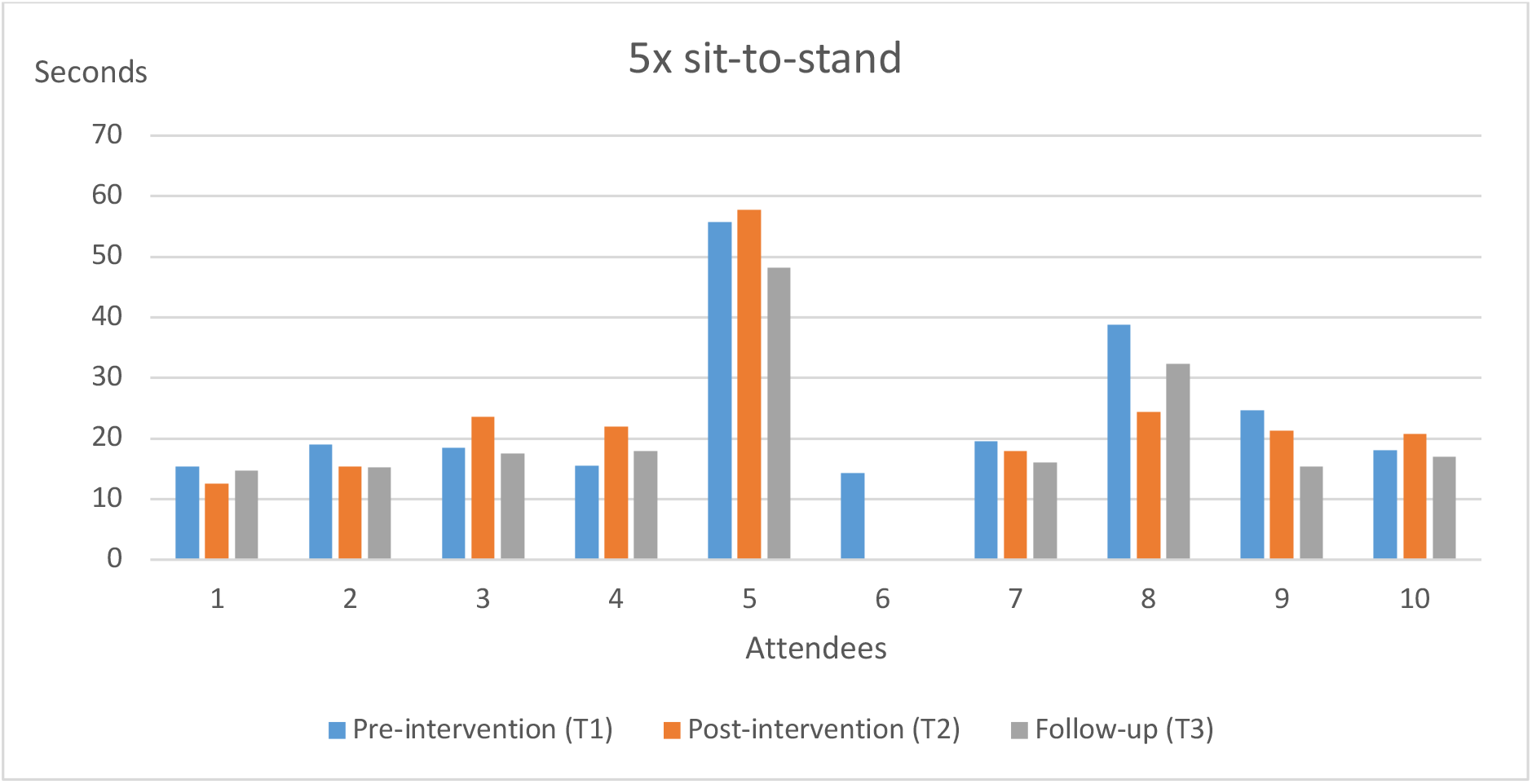
Evolution of the sit-to-stand test (expressed in number of seconds) at the 3 measurement moments.

**Figure 3.**
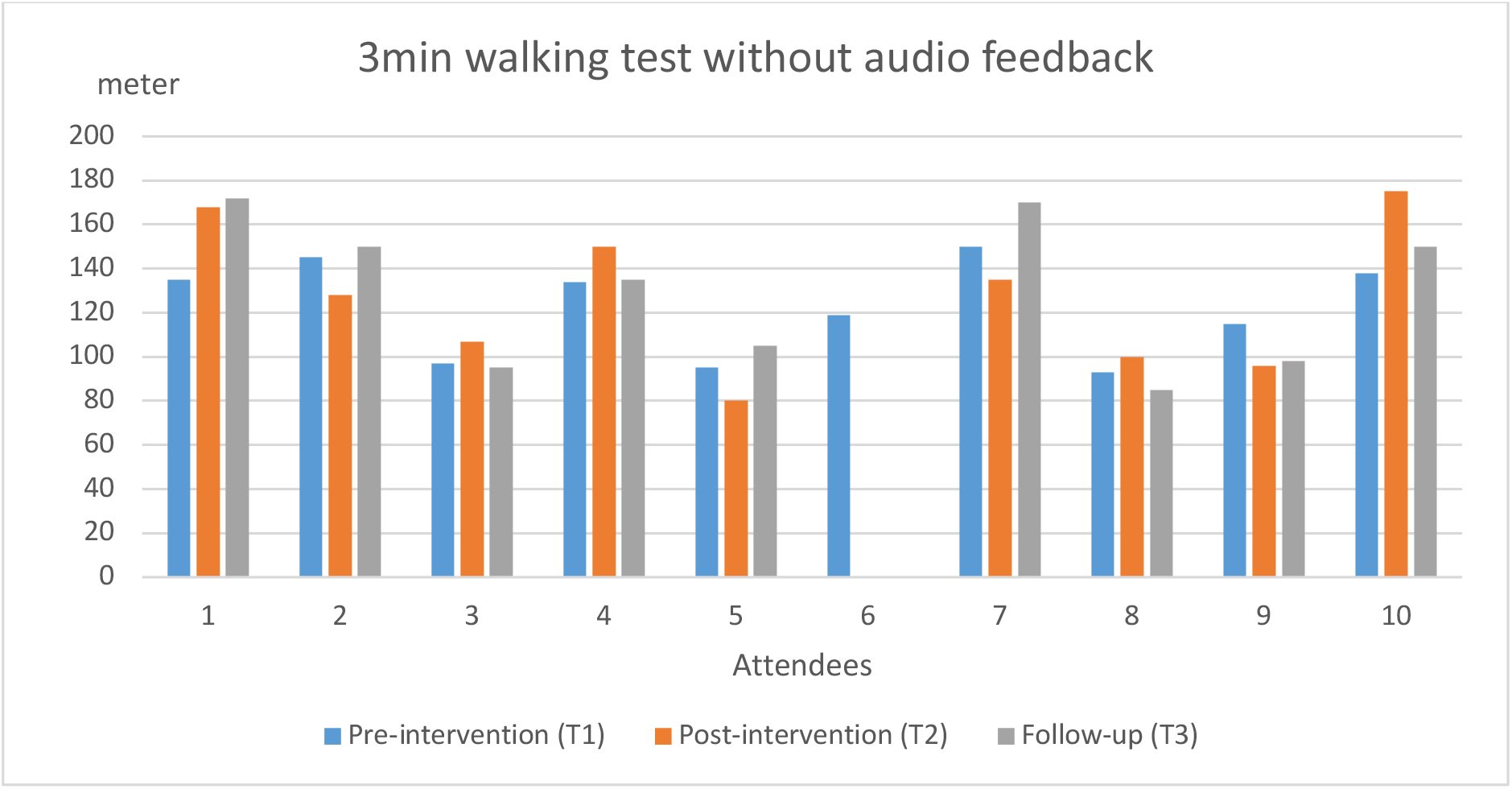
Evolution of the 3-minute walking test without audio feedback (expressed in number of meters covered in 3 minutes) at the 3 measurement moments.

Both figures show small differences between the different measurement moments. In the objective data no trend can be seen whether patients perform better or worse after the intervention period (large variability). At T1 and T2, the 3-minute walk test with audio feedback was also performed. The results of these measurements are shown in figure 4, figure 5 and figure 6.

**Figure 4.**
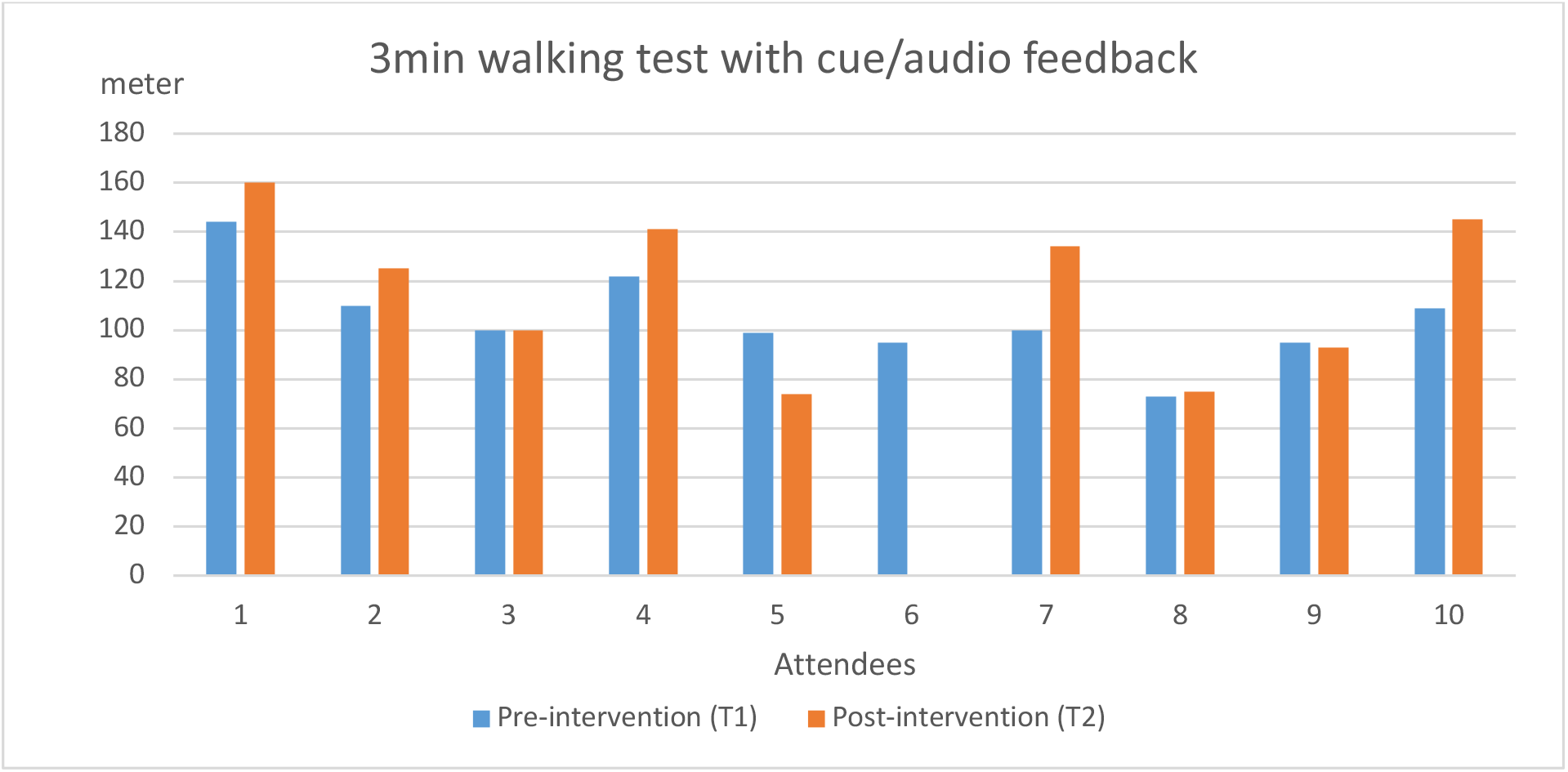
Evolution of the 3-minute walk test with audio feedback (expressed in number of meters covered in 3 minutes) at the 2 measurement moments.

**Figure 5.**
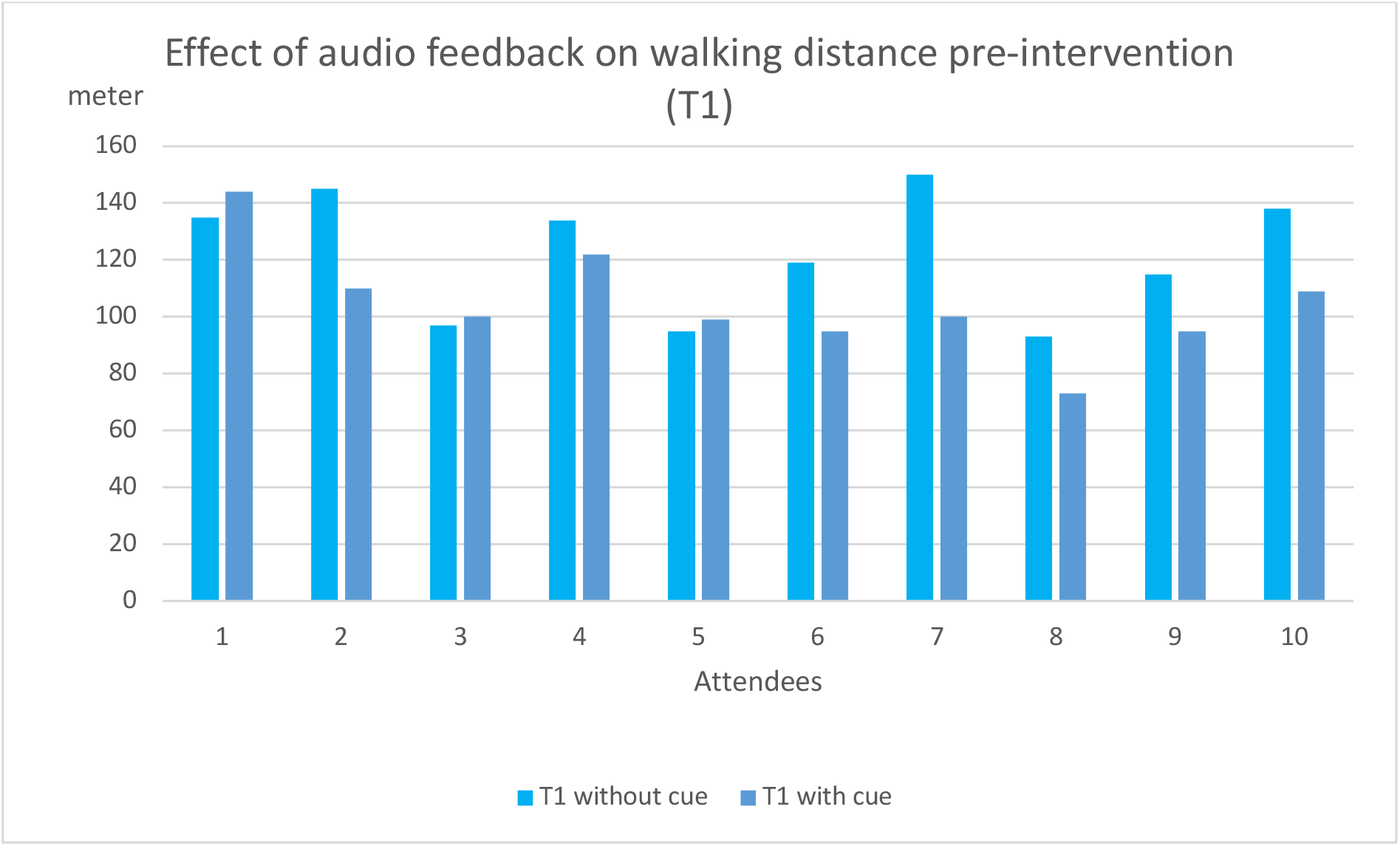
Effect of adding auditory feedback to the 3-minute walking test (expressed in number of meters walked in 3 minutes) at pre-intervention (T1)

**Figure 6.**
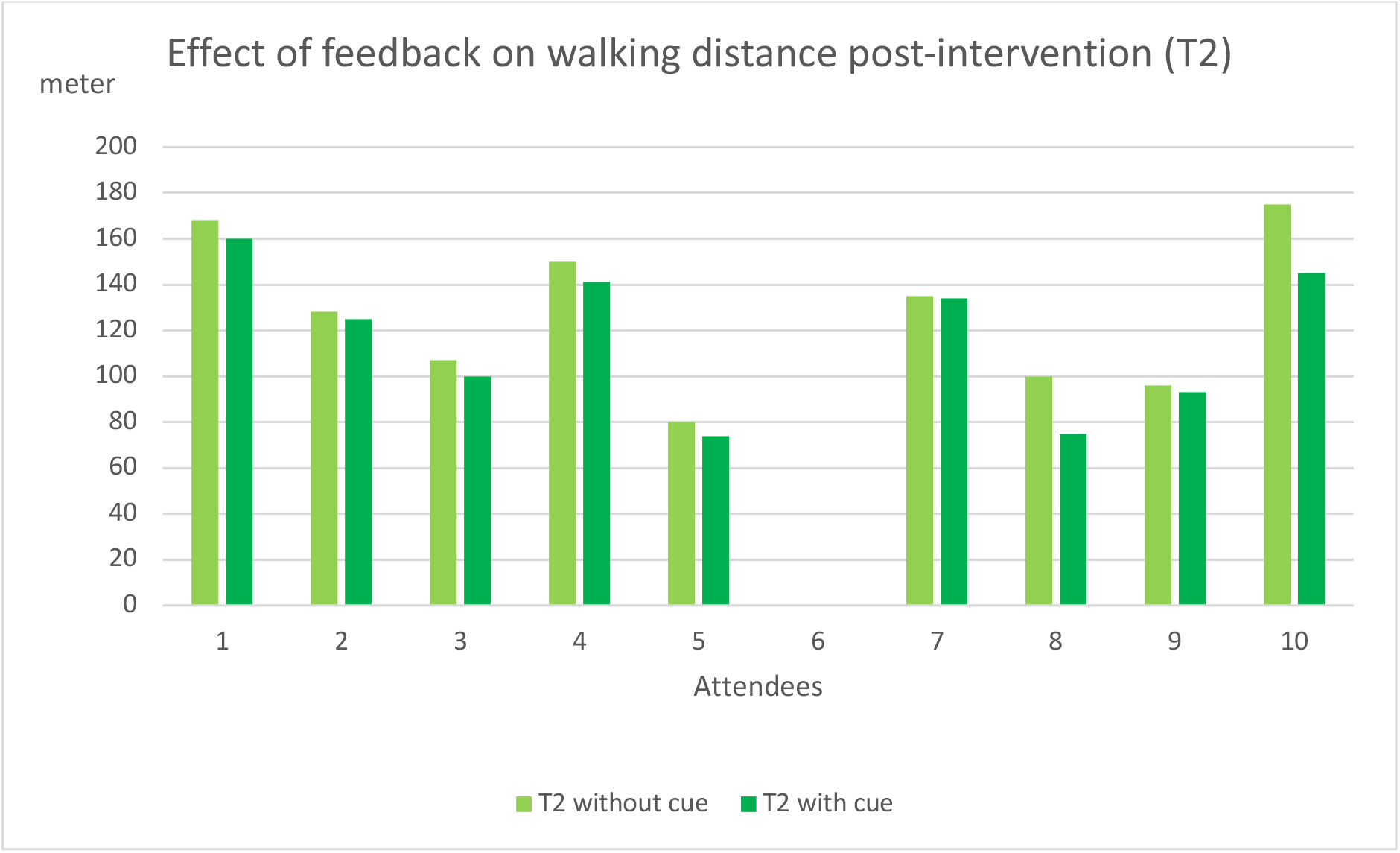
Effect of adding auditory feedback to 3-minute walking test (expressed in number of meters walked in 3 minutes) at pre-intervention (T2)

This figure shows a trend that patients at the post-intervention moment (T2) can usually cover a longer distance with the audio feedback compared to the pre-intervention measurement (T1) with audio feedback. This could indicate that due to the intensive training with Stride One, the patient has become more accustomed to its use and that this feedback can be processed better, leading to a better outcome. However, these results should be interpreted with caution, given the small sample size (and also some patients who do not show this trend).

Figure 5 and figure 6 show that adding auditory feedback has a negative influence on the number of meters the patient can walk at both measurement moments. However, this is only a decrease in the quantitative parameter. At each measurement with auditory feedback, a clear positive improvement was noticed by the therapist. This means that patients walk less distance, but are possibly more concentrated on a correct execution, searching for the positive feedback, which resulted in a qualitatively better roll-off pattern. This 1st primary endpoint is further discussed in chapter 5.

### 4.3 Experience & satisfaction of study participants

At the post-intervention measurement (T2), the satisfaction of the study participants was surveyed. The figure below shows the percentage of participants with the estimate of the general effect of Stride One on their gait pattern.

**Figure 7.**
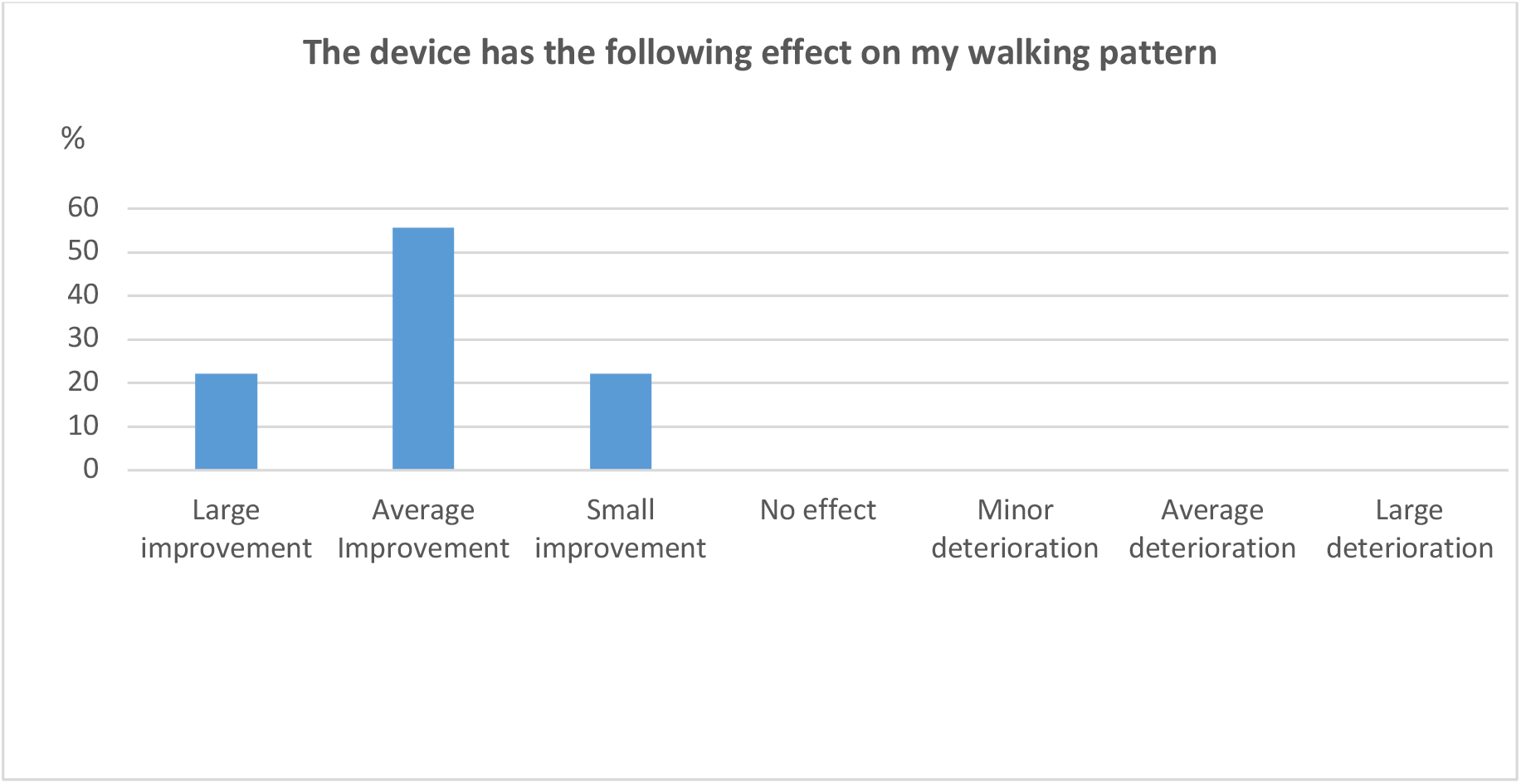
Estimated effect on gait pattern.

The table below shows for each statement how often this statement is true or false for the participants, and to what extent. It concerns absolute numbers of the participants who chose a category with the following score range: 0, completely false to 10, completely true.

**Table 2.** Satisfaction measured by different statements. Absolute numbers of patients who chose a certain category (from completely false to completely true) are shown (n=9)

|  | 0<br>Completely<br>false | 1 | 2 | 3 | 4 | 5 | 6 | 7 | 8 | 9 | 10<br>Completely<br>true |
| --- | --- | --- | --- | --- | --- | --- | --- | --- | --- | --- | --- |
| I feel anxious and embarrassed with the device | 6 | 2 |  |  |  | 1 |  |  |  |  |  |
| I feel tense with the device | 7 |  |  | 1 |  | 1 |  |  |  |  |  |
| I feel the device on my body | 6 |  | 1 | 1 |  | 1 |  |  |  |  |  |
| I feel the device moving or I feel that it is not properly attached | 8 |  |  |  |  | 1 |  |  |  |  |  |
| I have problems attaching or turning on the device | 7 |  |  |  |  | 1 |  |  | 1 |  |  |
| The device causes me pain | 9 |  |  |  |  |  |  |  |  |  |  |
| The device restricts my movements | 9 |  |  |  |  |  |  |  |  |  |  |
| I do not feel safe with the device | 9 |  |  |  |  |  |  |  |  |  |  |
| I feel that the device is not working properly | 2 |  | 1 |  |  | 5 |  | 1 |  |  |  |
| I experience using the sole as pleasant and enjoyable |  |  | 1 |  |  | 4 |  | 2 | 2 |  |  |
| I walk noticeably better when using the device |  |  |  |  | 1 |  | 1 | 4 | 2 |  | 1 |
| The device helps me understand better what the physiotherapist means |  |  | 1 |  |  |  | 1 | 3 | 1 |  | 3 |
| The device allows me to practice more and better |  |  | 1 |  |  | 2 | 1 | 2 | 1 |  | 2 |

The various statements generally show a great deal of satisfaction among the study participants. Patients are not worried or embarrassed about Stride One or generally do not feel tense with the device. It also does not cause them any discomfort when wearing Stride One, and they do not feel unsafe with the device. Several patients did indicate that the device does not always work properly. Most patients experience it as an added value; they feel that they walk noticeably better when using Stride One (100%), know better what their physiotherapist means (89%) and can practice more and better (67%).

## 5. Measuring gait quality with Stride One

The analyses and text of this chapter were added by Ceriter with the approval of Frame, the interpretation of the analysis was done together with Frame.

The first primary endpoint of the study is the complete and correct foot roll-off (detected and interpreted in the Ceriter platform). Stride One measures gait quality based on the parameters as mentioned under 3.4. For each step it measures whether a correct heel-strike, mid-stance and terminal stance are achieved. This results in a score (% correct) for each of the three parameters and an average score that shows the quality of roll-off, per step and averaged over all steps. The sensors in the sole are located at the places as indicated in the figure below:

**Figure 7.**
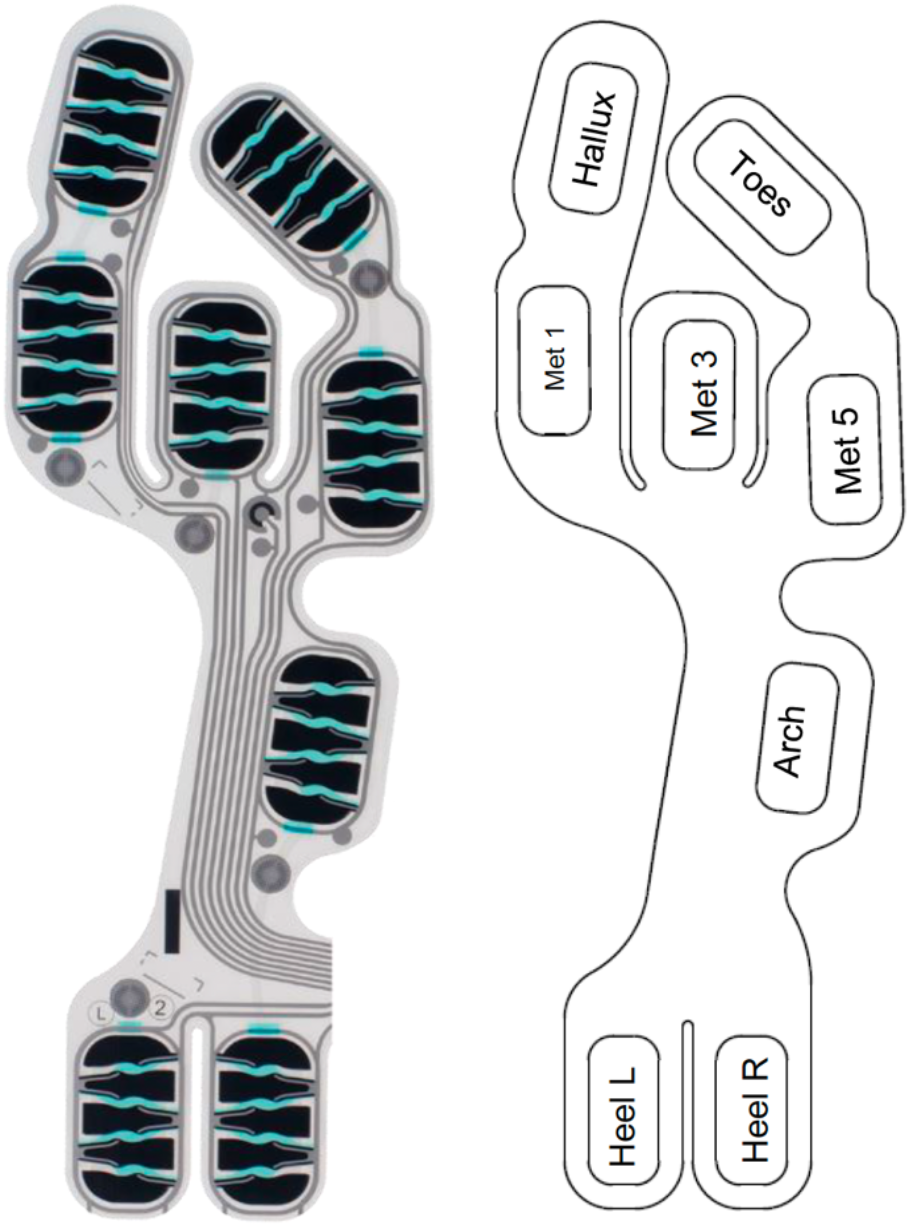
Location of the pressure sensors in the Ceriter sole.

A correct roll-off pattern looks like the figure below.

**Figure 8.**
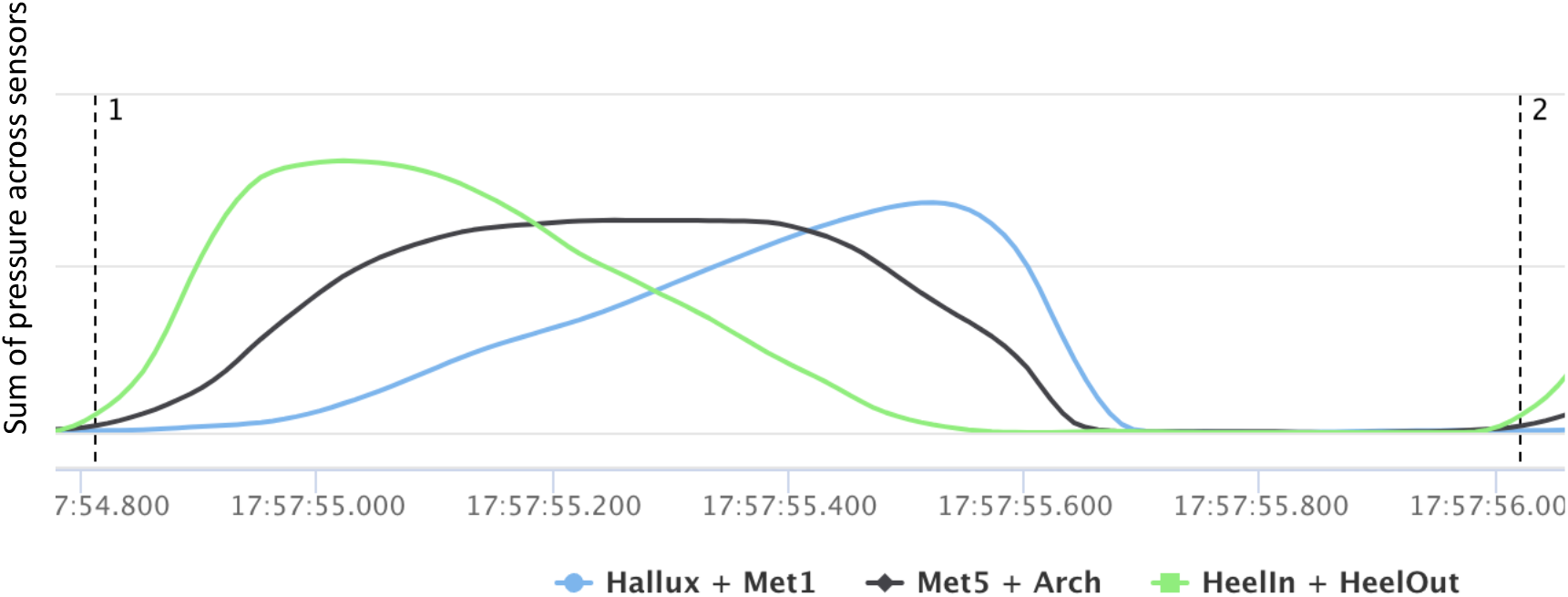
Correct roll-off pattern.

Heel-strike is considered correct if the sum of the 2 heel sensors reaches a maximum pressure at the beginning of the step. Mid-stance is considered correct if the sum of the Met 5 and Arch sensors reaches a maximum after a correct heel-strike. Terminal stance is considered correct if the sum of the Met 1 and Hallux sensors reaches a maximum at the end of the step.

### Average quality of the roll-off pattern

The following graphs show the impact on the average quality of the roll-off pattern for:

- Cue effect T1: the immediate impact of adding cues at time T1
- Cue effect T2: the impact of cues after a week of practice, at time T2
- Training effect: the impact on walking without cue after a week of practice (T2 compared to T1)
- Retention effect T3: the impact on walking without cue after 3 to 4 days without practice at time T3

**Figure 9.**
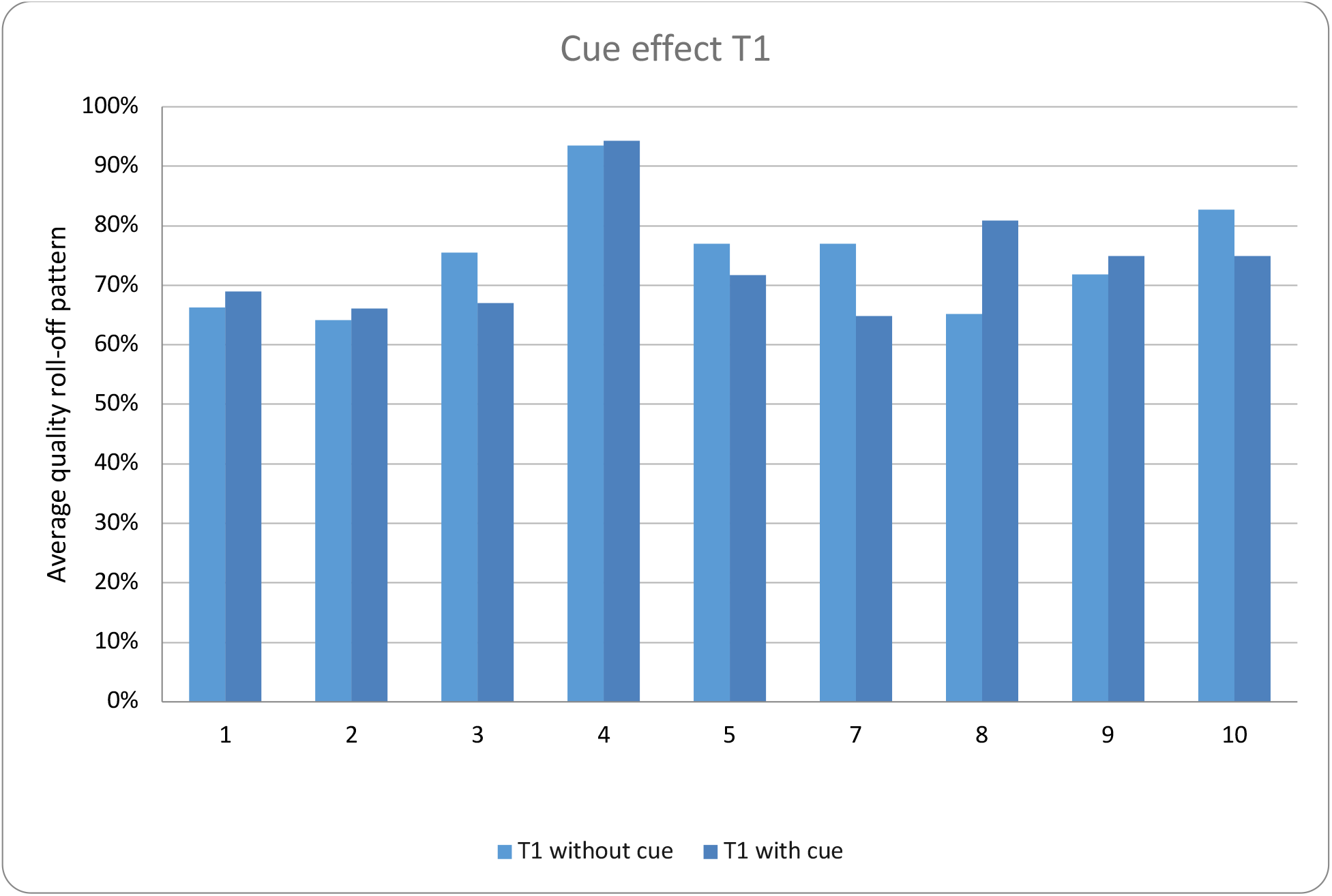
Cue effect T1, the immediate impact of cues at time T1.

The results indicate that there is no clear trend of the cue effect at time T1, even though therapists and patients report an improvement in gait.

**Figure 10.**
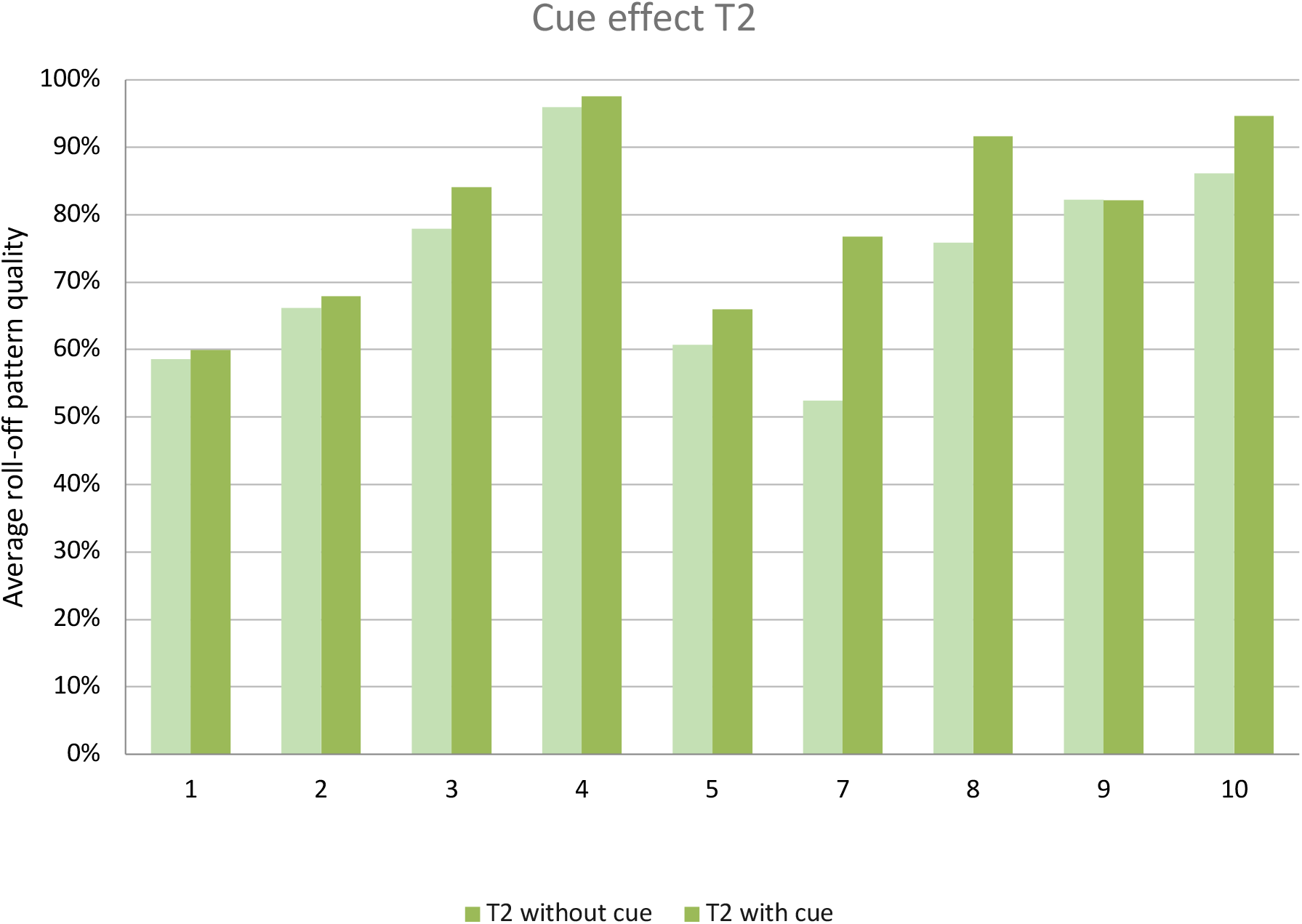
Cue effect T2, the impact of cues after a week of practice (at time T2)

The results of the test at time T2 show the quality of the roll-off pattern with cues is on average 8% higher than the one without cues after one week of practice with Stride One. Both these results and the feedback from patients and physiotherapists show an improvement in 100% of the cases, indicating a clear, clinically relevant impact.

There is no clear trend for training (Figure 11) and retention effect (Figure 12) on the roll-off pattern without cues. This is in line with the expectation that offering specific gait therapy should last 4 to 6 weeks for a good and lasting improvement to occur.

**Figure 11.**
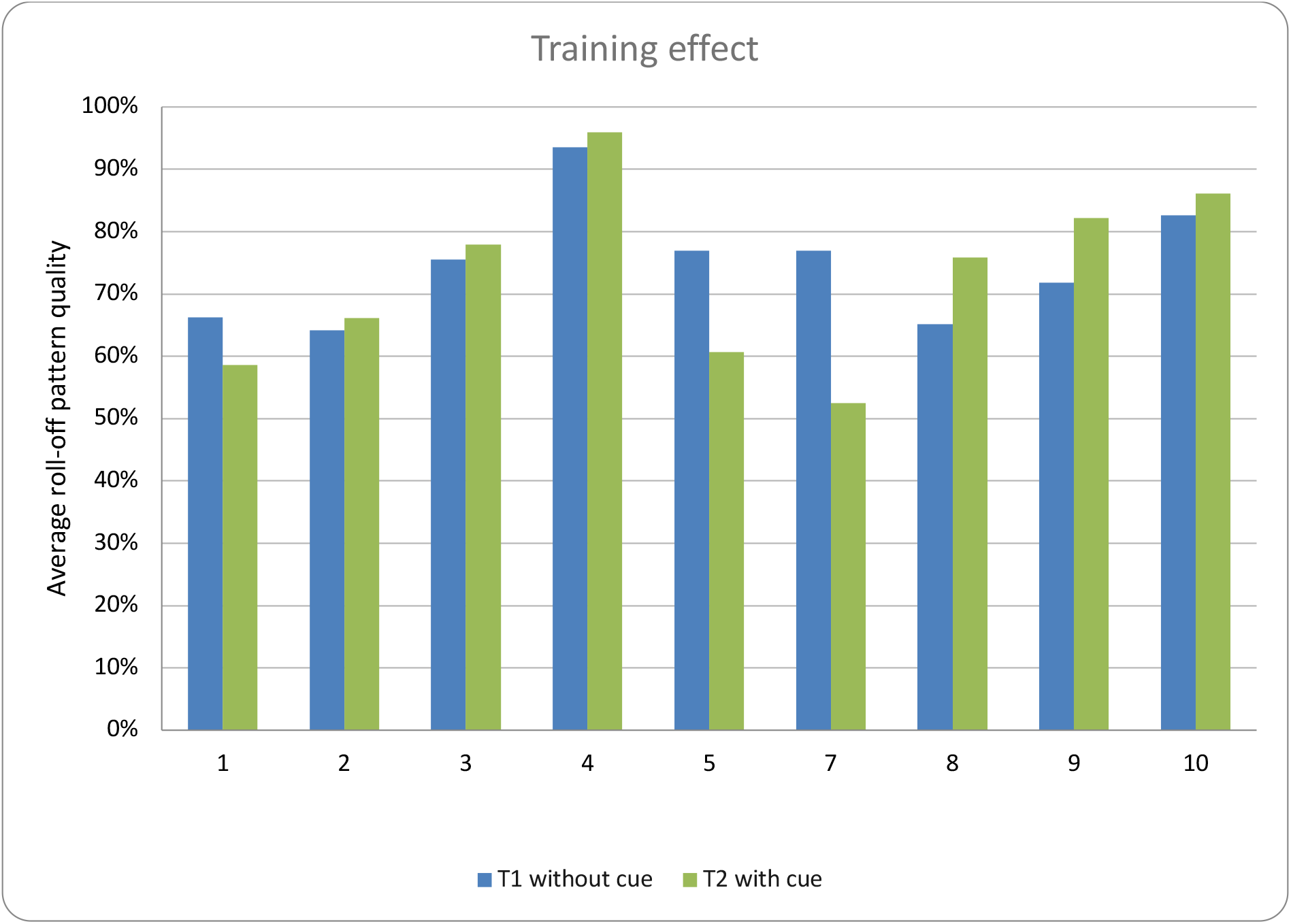
Training effect, the impact of steps without cue after one week of practice (T2 compared to T1)

**Figure 12.**
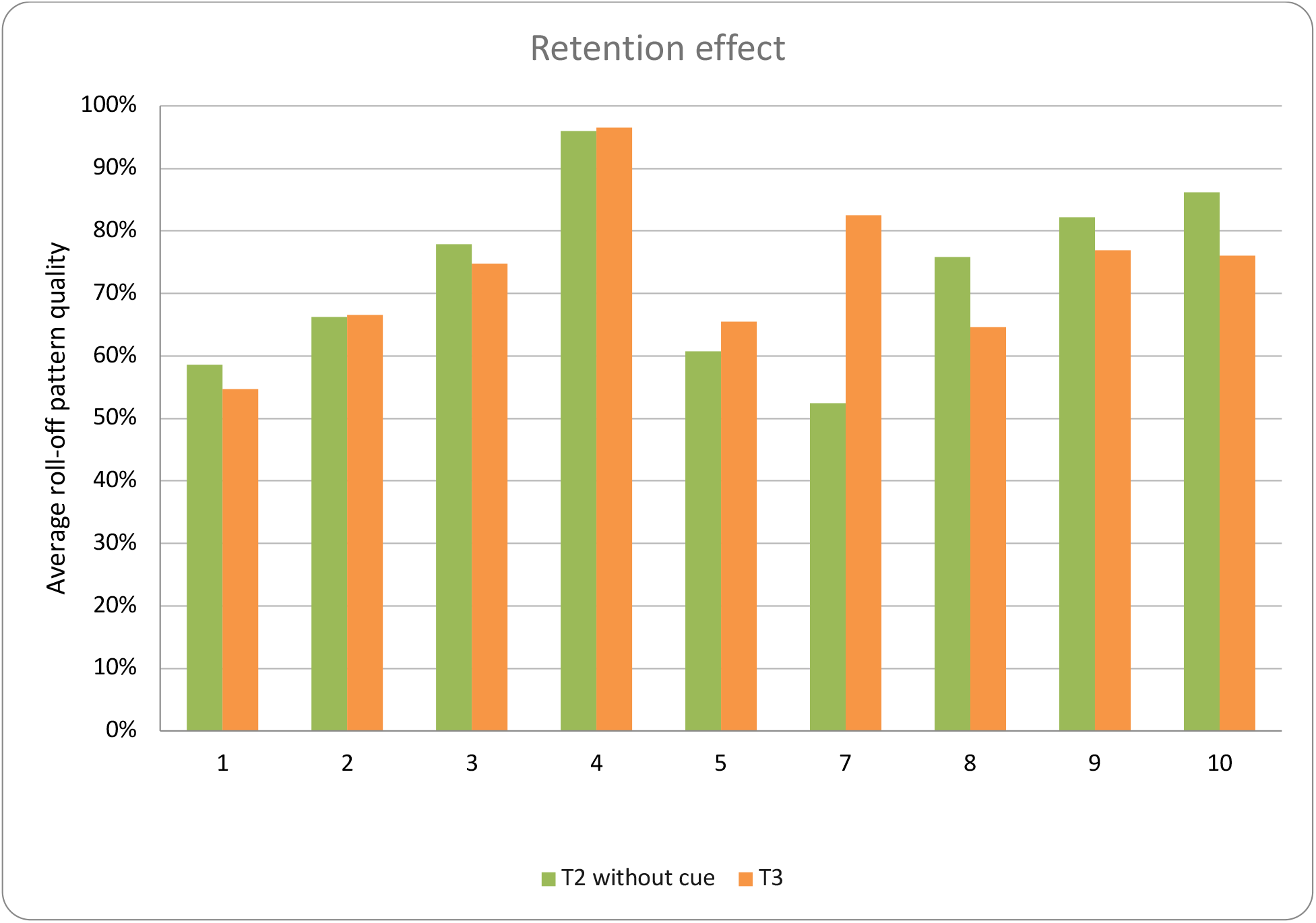
Retention effect, the impact of steps without cue after 3 to 4 days without practice at time T3.

**Figure 13.**
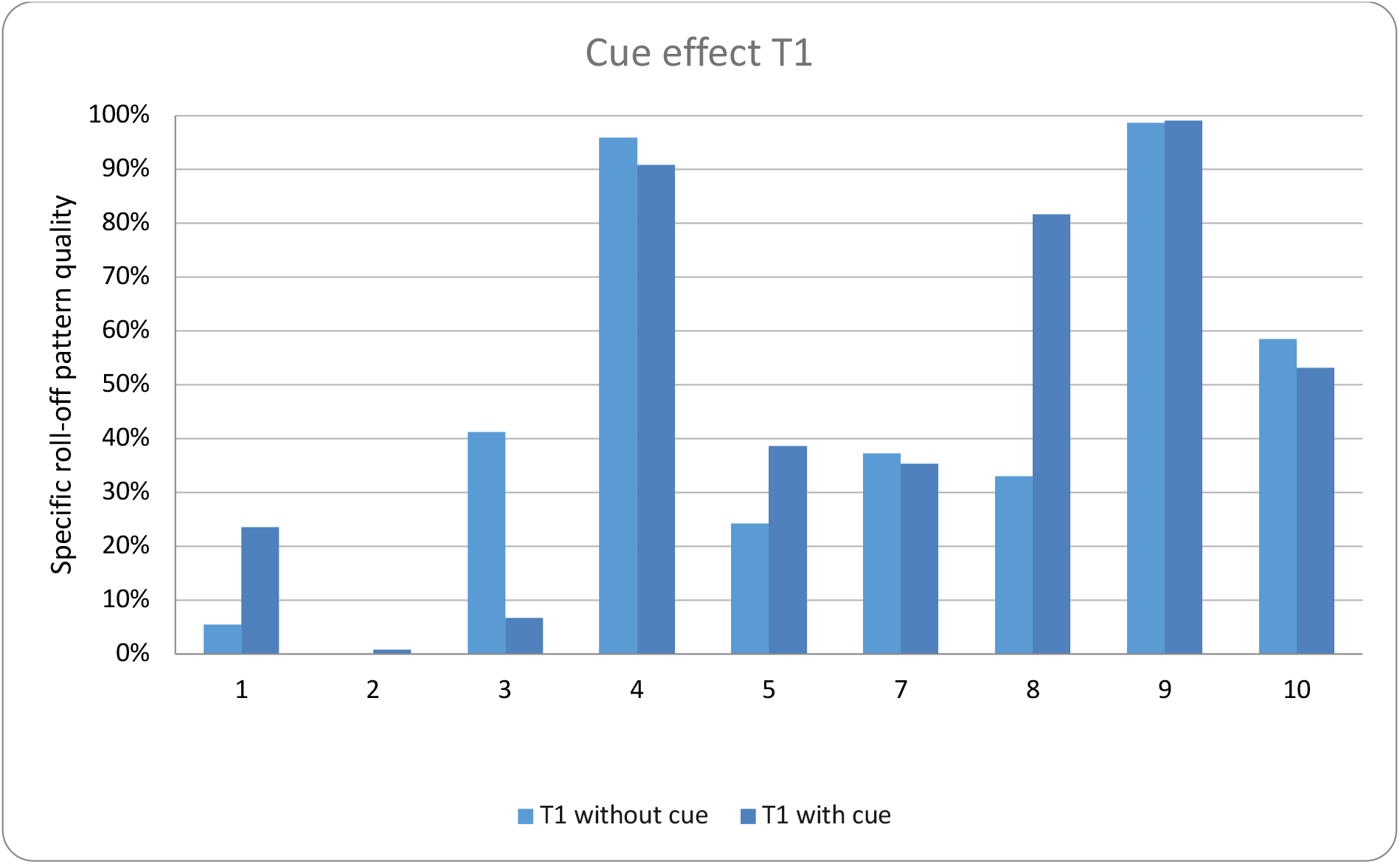
Specific cue effect T1, the immediate impact of cues at time T1.

**Figure 14.**
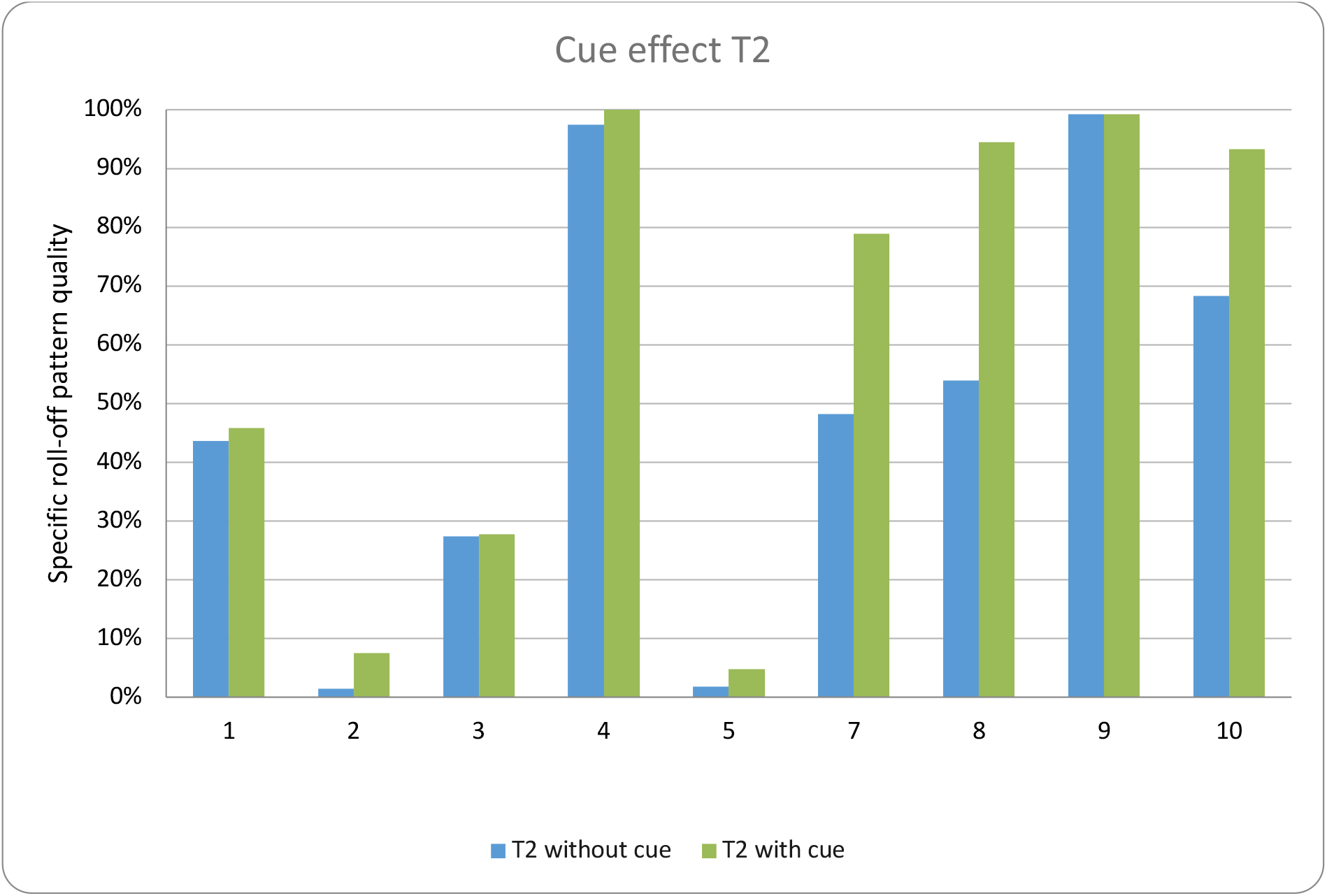
Specific cue effect T2, the impact of cues after one week of practice (at time T2.

**Figure 15.**
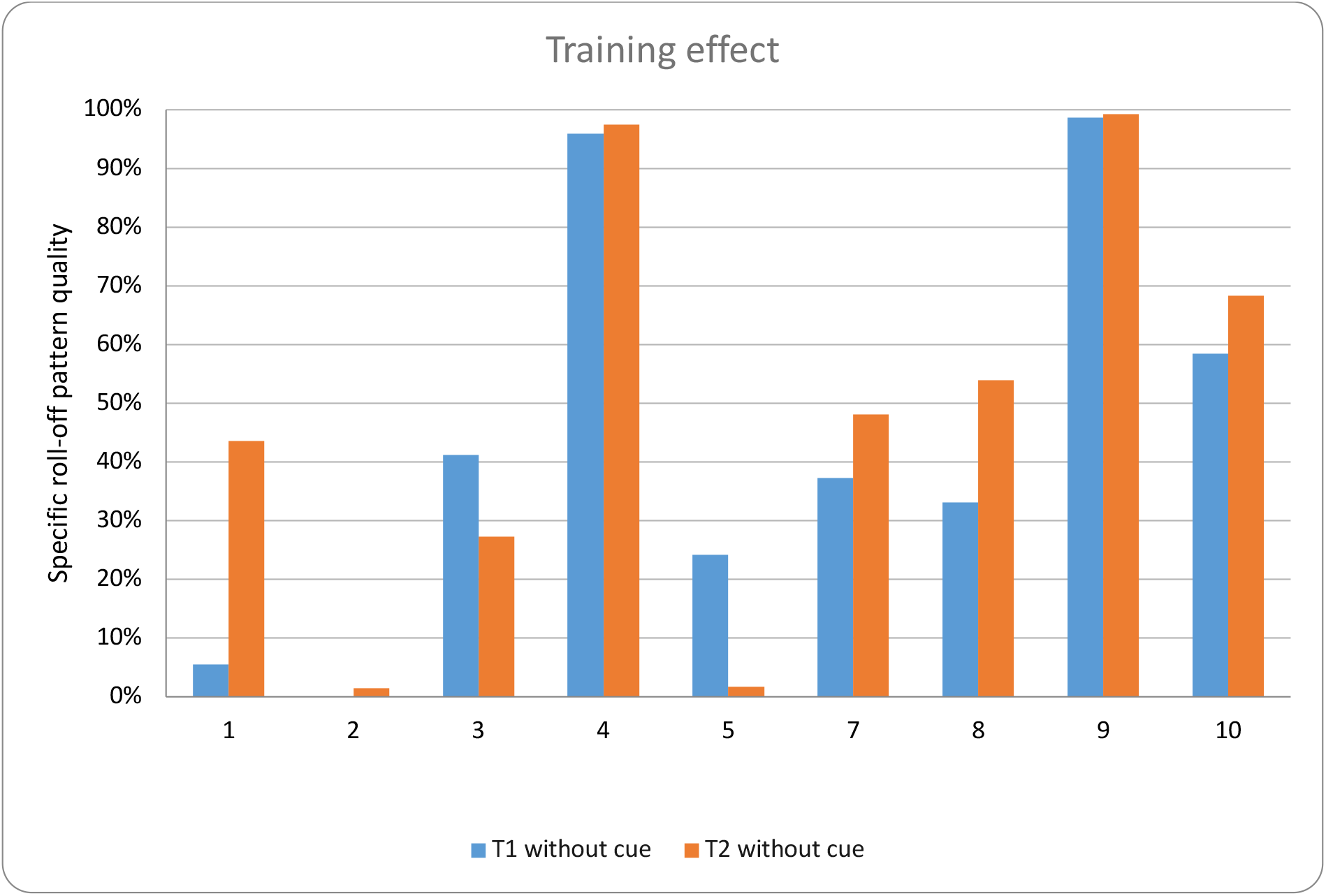
Specific training effect, the impact of steps without cue after one week of practice (T2 compared to T1))

**Figure 16.**
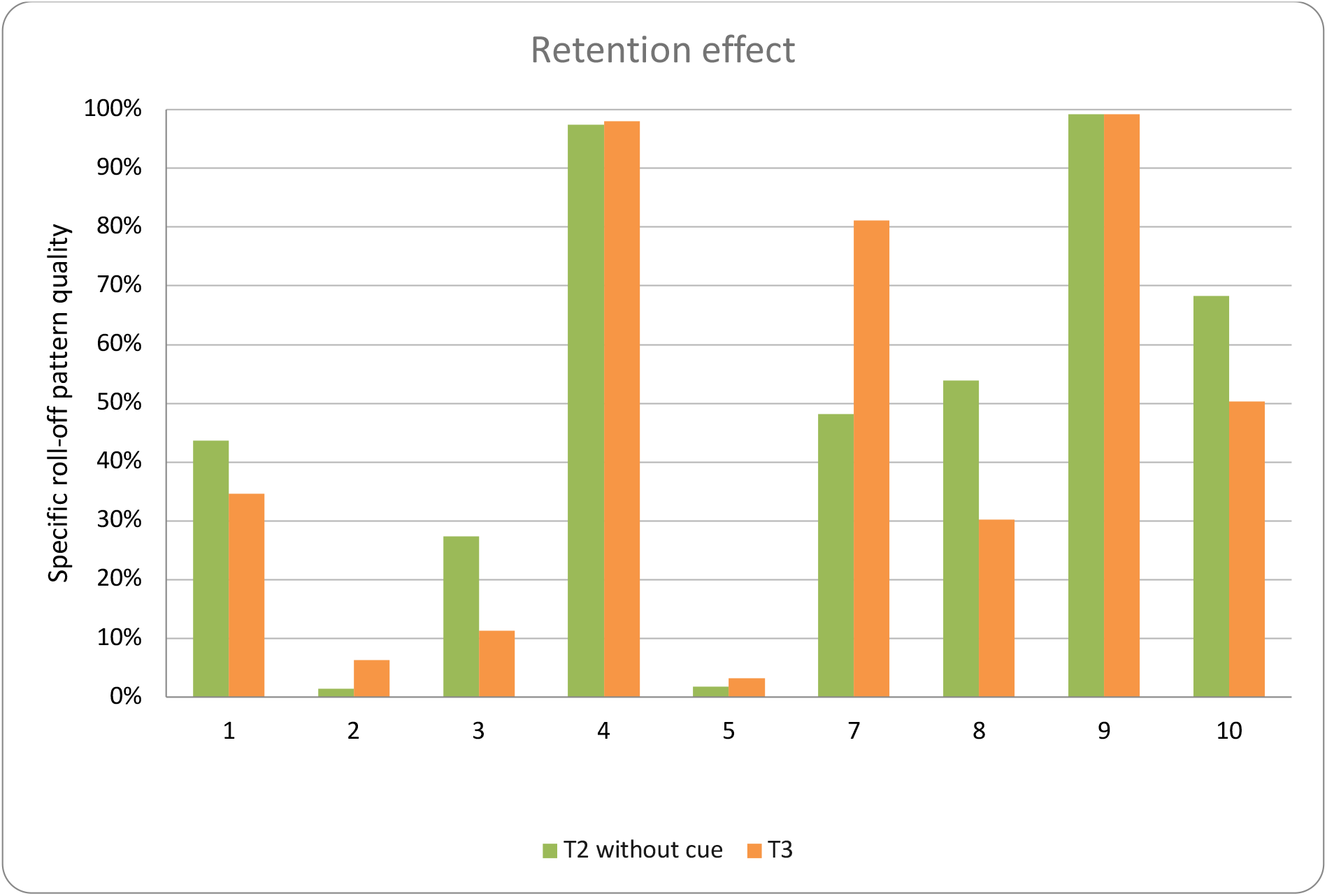
Specific retention effect, the impact of steps without cue after 3 to 4 days without practice at time T3.

### Specific quality of the roll-off pattern

Practicing with cue focuses on improving one specific aspect or parameter of the roll-of pattern (for example correct heal-strike). The impact of practicing was also calculated for the quality of the specific parameter that was practiced on by the respective patients.

Analysis of the graphs below confirms the conclusions that were drawn based on the evolution of the general roll-of pattern. There is a clear improvement of the specific roll-of quality after one week of practice of 25% on average. However, the practice period is too short to be able to speak of a training or retention effect when walking without cue.

## 6. Further research questions

Building upon this study’s already demonstrated clinically relevant impact, several questions remain open and can be investigated in a subsequent study:

1. What training period is needed to achieve a training effect, namely an improvement in the roll-off pattern when Stride One is not used?
2. How strong is the retention of the improvement in the roll-off pattern when Stride One is not used for a longer period of time?
3. Are there demographic or other characteristics that distinguish possible responders from non-responders?
4. Is there a correlation between the improvement in the roll-off quality and other parameters such as the number of meters covered during a walking test?

## Data Availability

All data produced in the present study are available upon reasonable request to the authors

